# Integrated Vaccination and Non-Pharmaceutical Interventions based Strategies in Ontario, Canada, as a Case Study: a Mathematical Modeling Study

**DOI:** 10.1101/2021.01.06.21249272

**Authors:** Matthew Betti, Nicola Luigi Bragazzi, Jane Heffernan, Jude Kong, Angie Raad

## Abstract

**Background:** Recently, two “Coronavirus disease 2019” (COVID-19) vaccine products have been authorized in Canada. It is of crucial importance to model an integrated/combined package of non-pharmaceutical (physical/social distancing) and pharmaceutical (immunization) public health control measures.

**Methods:** A modified epidemiological, compartmental SIR model was utilized and fit to the cumulative COVID-19 case data for the province of Ontario, Canada, from September 8, 2020 to December 8, 2020. Different vaccine roll-out strategies were simulated until 75 percent of the population is vaccinated, including a no-vaccination scenario. We compete these vaccination strategies with relaxation of non-pharmaceutical interventions. Non-pharmaceutical interventions were supposed to remain enforced and began to be relaxed on either January 31, March 31, or May 1, 2021.

**Results:** Based on projections from the data and long-term extrapolation of scenarios, relaxing the public health measures implemented by re-opening too early would cause any benefits of vaccination to be lost by increasing case numbers, increasing the effective reproduction number above 1 and thus increasing the risk of localized outbreaks. If relaxation is, instead, delayed and 75 percent of the Ontarian population gets vaccinated by the end of the year, re-opening can occur with very little risk.

**Interpretation:** Relaxing non-pharmaceutical interventions by re-opening and vaccine deployment is a careful balancing act. Our combination of model projections from data and simulation of different strategies and scenarios, can equip local public health decision- and policy-makers with projections concerning the COVID-19 epidemiological trend, helping them in the decision-making process.

## 1 Introduction

The “Severe Acute Respiratory Syndrome-related Coronavirus type 2” (SARS-CoV-2) is a novel, emerging coronavirus, belonging to the class of enveloped, single-stranded, positive-sense RNA viruses [1]. SARS-CoV-2 is the infectious agent responsible for the still ongoing “Coronavirus disease 2019” (COVID-19) pandemic. This respiratory disease is generally mild and even asymptomatic, but in a proportion of cases can worsen, becoming particularly severe and even life-threatening [2].

Due to the lack of effective vaccines and drugs that could prevent the infection and properly treat patients, as well as given the highly contagious nature of the virus and its quick spread, public health agencies have decided to implement non-pharmaceutical interventions (NPIs) [3]. NPIs have become necessary also due to the unprecedented strain on the health systems worldwide. Two major strategies have been developed: an eradication and a mitigation strategy, based on the stringency of the measures adopted [3].

Several vaccine candidates have been designed, and, recently, some of them have entered phase III clinical trials [4]. Immunization campaigns are anticipated to be particularly challenging in that a large number of subjects needs to be vaccinated in a short span of time. Countries will have to face unprecedented logistic and organizational issues, coping with shortage of health-care personnel staffing. It can be expected that, before immunizing an adequate amount of individuals necessary to achieve herd immunity, public health authorities will have to continue enforcing NPIs. Mathematical modelling can assist public health decision- and policy-makers in the decision-making process, regarding the dynamical modulation of the de-escalation/relaxation of the public health measures adopted, towards a gradual re-opening strategy. We will take Ontario, Canada, as a case study, but our model is a general-purpose one and, after being properly parameterized and informed with locally available data, can be adapted to any country/scenario.

### 1.1 Non-Pharmaceutical Interventions in Ontario

Canada belongs to those countries which have decided to implement a mitigation rather than an eradication strategy. The first confirmed COVID-19 case was reported on January 25th, 2020 and, since then, as of January 2nd, 2020, Canada has experienced a burden of 590,280 cases, with 15,715 deaths. The Federal government and the province of Ontario have enforced an integrated package of public health interventions. In particular, the Ontario province has closed schools and universities on March 14, declared emergency on March 17, restricting public events and recreational venues, and closed non-essential workplaces on March 24. These control measures have been escalated and gradually become more and more stringent, significantly contributing to mitigate against COVID-19 and saving lives.

### 1.2 Vaccine Deployment in Ontario

Currently, two mRNA-based COVID-19 vaccines have been authorized by the Canadian regulatory body “Health Canada”.

On December 14, 2020, Canada started administering the first vaccine doses, according to the federal vaccine plan shown in Figure 1. The Ontario Government plan consists of three phases: during the phase I, vaccine doses are limited and healthcare workers working in hospitals, long-term care and retirement homes, congregate care settings, and other health facilities as well as members of remote, under-served, either urban or rural, Indigenous communities (such as First Nation communities, Métis and Inuit adults) are being prioritized. During phase II, which is likely to start in Winter 2021, the list of people who can get vaccinated will be further expanded. Finally, during phase III, any person willing to be immunized will receive its vaccine doses. Specifically concerning the Ontario province, more than 20 hospitals will be involved in the immunization campaign.

**Figure 1:**
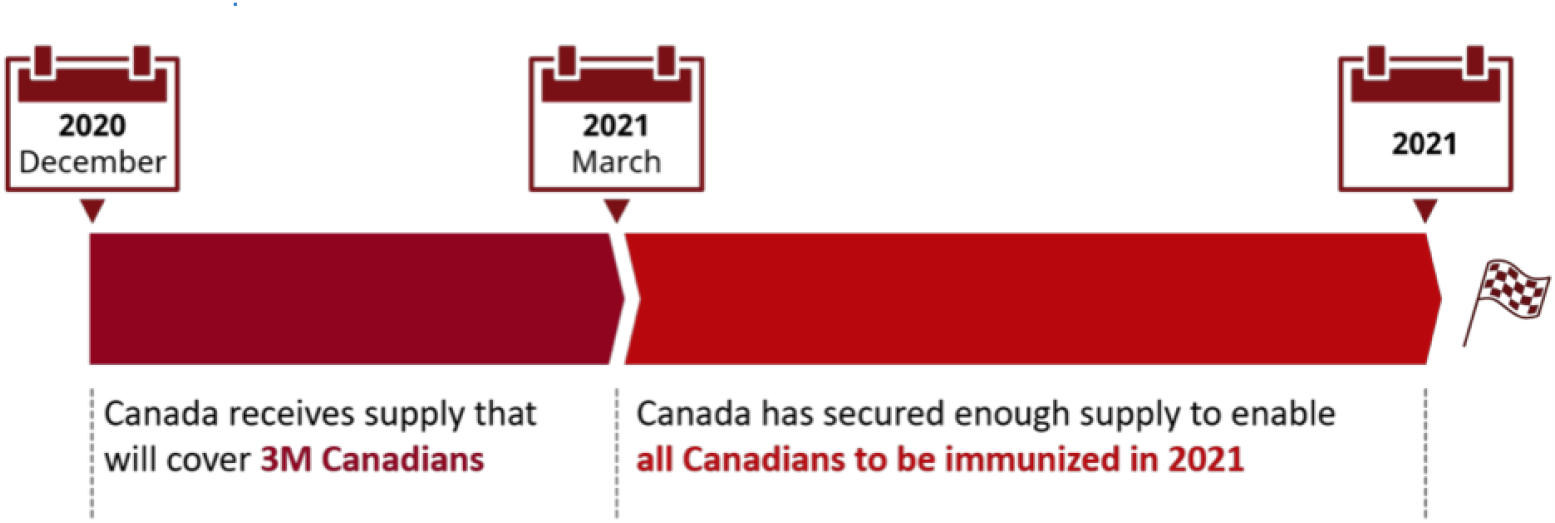
The Federal Government vaccine roll-out plan, adapted from [5].

## 2 Methods

### 2.1 COVID-19 dynamics model

We use the model and fitting techniques developed in [6]. The modified SIR model is developed to forecast epidemic trajectories using minimal cumulative case data of an outbreak. The model in [6] is used to estimate and quantify the rate of under-reporting, the efficacy of preventative measures in healthcare settings, the basic reproduction number, and the efficacy of NPIs. The model uses re-scaled time; time is measured in infection incubation periods, 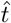 to further reduce the required parameters.

The compartments we are interested in are the number of mild cases, *I*_*m*_, the number of severe cases, *I*_*s*_, the cumulative known cases *C*_*K*_ and cumulative incidence *C*_*I*_ We make the following assumptions in the model:

- The total population is constant.
- Acquired immunity lasts longer than the outbreak.
- There is no co-infection or super-infection.
- The testing/reporting rate in the population is relatively constant.
- The probability of a case being severe vs. mild is constant.
- All severe infections are reported, whereas a fraction of mild infections are.

The model equations are then given as

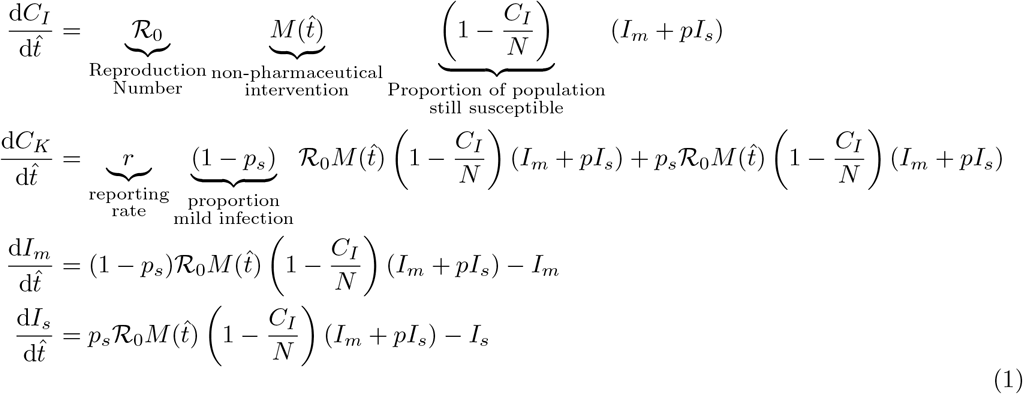

where *N* is the total susceptible population, ℛ_0_ is the basic reproduction number, *p*_*s*_ is the probability of severe infection, *r* is the average testing/reporting rate, 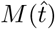 is a mitigation function, and 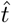 is time scaled by the average infectious lifetime 1*/µ*. Model (1) can be rewritten as:

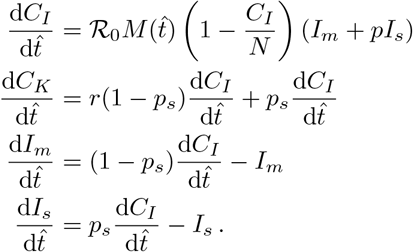

Note that Model (1) accounts for, but does not assume, that severe infections are less infectious than mild or asymptomatic infections by a factor of *p*. We include *p* since in novel or dangerous epidemics severe cases are often managed and quarantined once known.

Model (1) is coupled with four initial conditions, derived from three parameters:

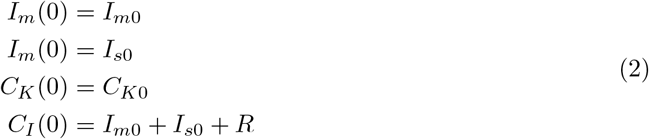

where *R* is the number of cases that have already recovered or died by the time we start fitting data.

The function *M* (*t*) is a predetermined function that captures the effects of non-pharmaceutical interventions on rates of infection. When fitting, we use a functional form of

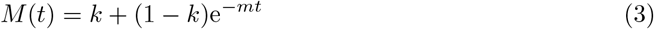

This allows us to quantify both socially-driven relaxation and adherence to non-pharmaceutical interventions.

### 2.2 Vaccine Deployment vs. Social Distancing

The primary aim of vaccination is to reduce the number of susceptible individuals in the community. This is accounted for with the change *N → N* (*t*). *N* (*t*) is defined as a function which starts at *N* (*ŝ*) = *N* where *N* is the total population and ŝ is the day of vaccine deployment, and approaches the expected vaccinated population *N* (*s*^∗^) = *vN* for 0 *≤ v ≤* 1.

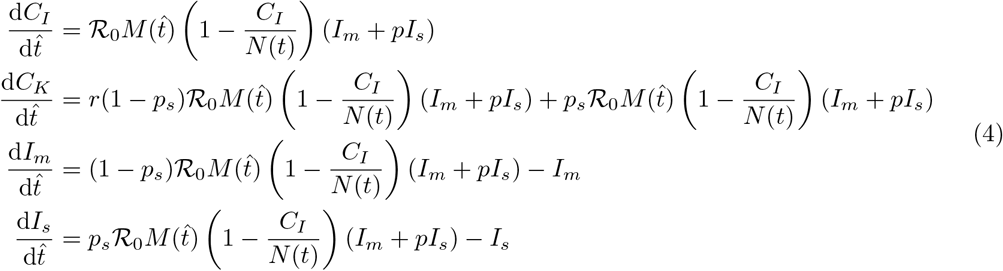

As an example, we employ

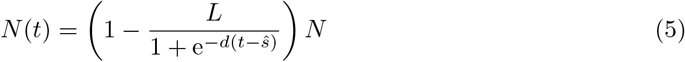

Where *L* is the proportion of the population we expect to be vaccinated, *s*^∗^ is as above and *d* is a shape parameter that determines how quickly the vaccine is deployed.

### 2.3 Data Fitting

We fit our model to the cumulative case data for the province of Ontario from September 8, 2020 to December 8, 2020 for the 2020 COVID-19 pandemic. September 8, 2020 is chosen because of the observed significant increase in the number of reported infected cases Fits were done until December 8, 2020 as at the time of the analyses this was the most current data available. Of course, projections can be adjusted by taking more recent data as it is made available.

To reduce the number of scenarios considered, we fit system (1) to cumulative case data prior to vaccine deployment. Using the fitted parameters, we can then explore different functional forms of *N* (*t*) to determine the effects of different vaccination strategies on the epidemic curve. We fit the vector of parameters:

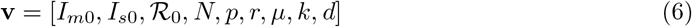

using least-squares on the cumulative reported cases per day *and* the new daily reported cases

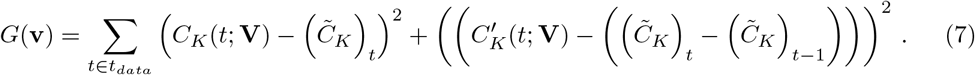

#### 2.3.1 Outcome of different vaccine roll-out strategies

Using the estimated parameters we explore different scenarios that could result with vaccine roll-out. We start by defining a logistic roll-out strategy

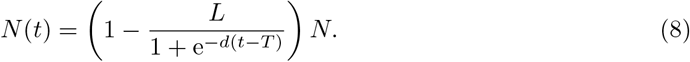

Using public data about the vaccine roll-out [5, 7], we set parameters of *N* (*t*). We use *L* = 0.75 based on a STATCAN survey [7]; meaning we are limited to 75% of the population of Ontario getting vaccinated. Using the projections from [5], we set *d* and *T* such that approximately 10% of the population will be vaccinated by March 2021 and all voluntary vaccination will be completed by the end of 2021. This gives us *d ≈* 0.433 and *T* = 24.73, keeping in mind our re-scaled time from the fitting. We first study the outcome of this vaccination strategy assuming that the non-pharmaceutical interventions are to remain in place at the current levels of acceptance and adherence. These parameters are summarized in Table 1 and the vaccination curve is shown in Figure 2.

**Table 1:**
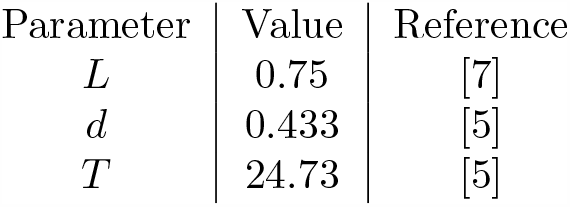
Parameter values to inform equation (8). Values are taken from literature to best match the current public sentiment of a COVID-19 vaccine and its the government’s plans to roll-out the vaccine.

| Parameter | Value | Reference |
| --- | --- | --- |
| $L$ | 0.75 | [7] |
| $d$ | 0.433 | [5] |
| $T$ | 24.73 | [5] |

**Figure 2:**
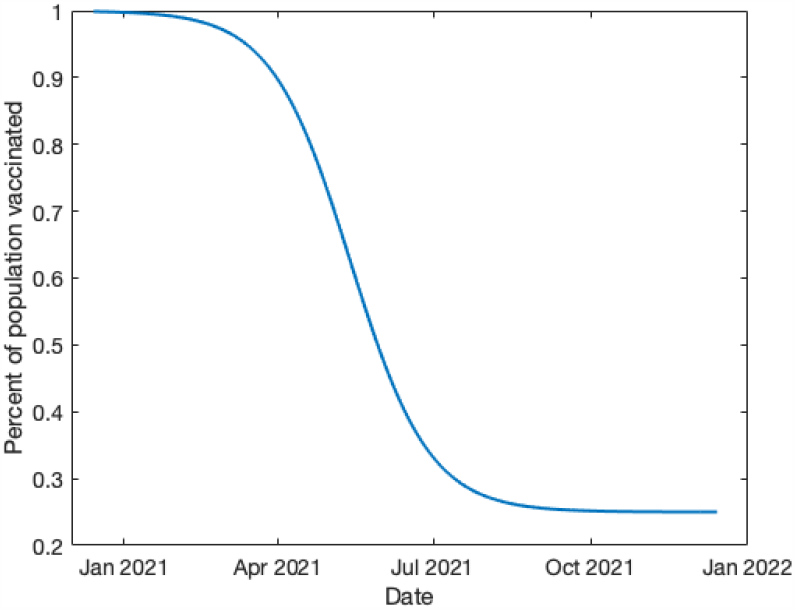
A visualization of equation (8). We can see that by March 2021, 10% of the population of Ontario is expected to be vaccinated. By the end of 2021, we expect to approach 75% of the population to be vaccinated.

Of course, at some point non-pharmaceutical interventions must be lifted. We can compete these two factors by modifying *M* (*t*). If we allow

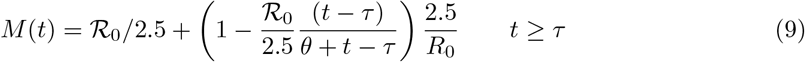

for *t* > *t*^∗^ where *t*^∗^ is when we begin relaxation of non-pharmaceutical interventions. We set *τ* to be the date relaxation begins and set *θ* accordingly so that by the end of 2021, non-pharmaceutical interventions are completely relaxed. We maintain that *M* (*t*) is a function of time alone so that we are can remain general about how and when these relaxations may manifest themselves. For instance, non-pharmaceutical interventions may relax through policy, fatigue, or apathy as interventions remain in place for long periods of time.

## 3 Results

### 3.1 Parameter estimation

Table 2 details the estimated parameter values obtained from fitting the vaccine and social dynamics model to the reported cumulative case data for the province of Ontario from September 8 to December 8, 2020. As expected, the estimated ℛ_0_ is below the current accepted basic reproduction number of ℛ_0_ = 2.5 [6, 8, 9]; this is a result of using the reported data from September 2020, where NPIs are in place.

**Table 2:** Estimated model parameters for Ontario for September 8, 2020 to December 8, 2020

|  |  |
| --- | --- |
| $\hat{t}$ | $t/10$ |
| $\mathcal{R}_0$ | $1.58 \pm 0.16$ |
| $p$ | $0.0018 \pm 0.01$ |
| $r$ | $0.0004 \pm 0.002$ |
| $k$ | $0.81 \pm 0.27$ |
| $m$ | $0.54 \pm 0.41$ |

### 3.2 Outcome of different vaccine roll-out strategies

#### 3.2.1 Assuming a 75% vaccination rate in Ontario with fixed NPIs

Shown in Figure 3 are the total cases (purple), known cases (green), active mild cases (red) and active severe cases (blue). We see that all active cases are resolved by September 2021, given the vaccination strategy used in the simulation.

**Figure 3:**
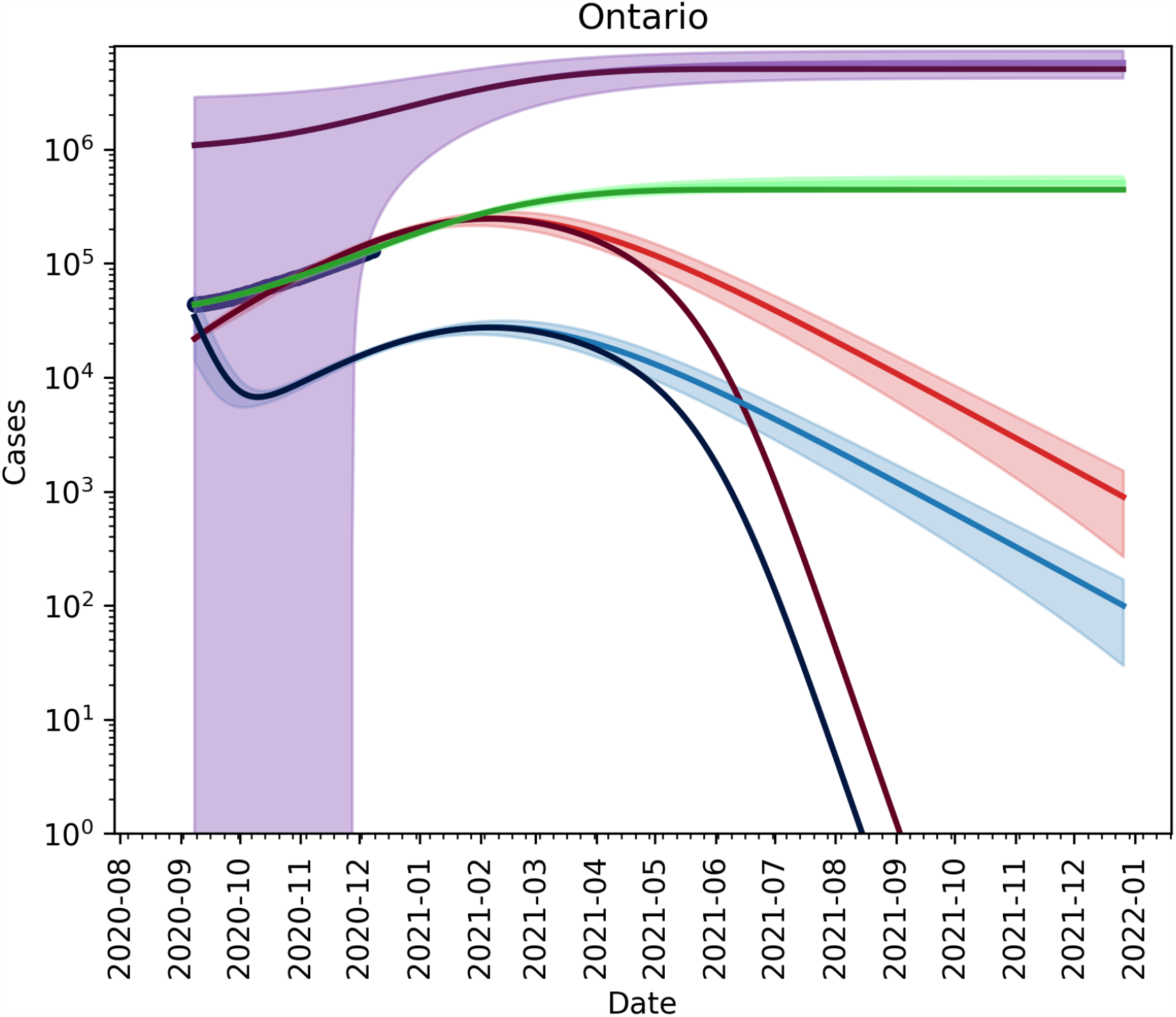
Light lines: No vaccine trajectories. Dark lines: With Vaccine using (8). Shaded areas represent the 95% confidence interval obtained from the fits. The current vaccination roll-out plan will not take hold in the population until *after* we reach the peak of the outbreak.

As shown in Figure 4, when considering the current infection rate, with the assumption that will approach 75% vaccination of the population by September 2021, we observe that the effect of the vaccine will only be seen after the peak has occurred where the province will be able to ensure that the outbreak end around July 2020 (pink line) rather than January 2022, as seen in the case of no vaccine roll-out (blue line).

**Figure 4:**
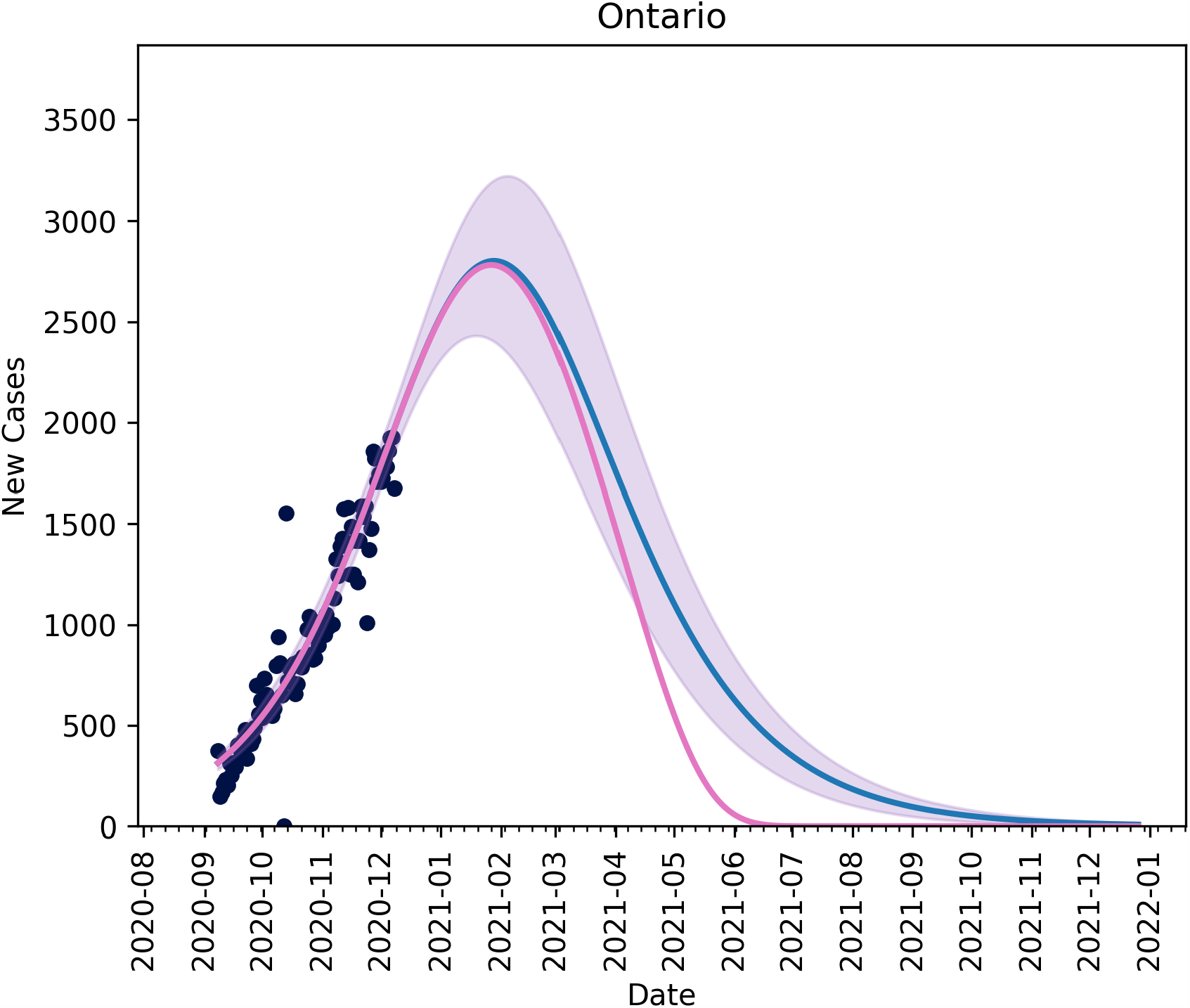
The blue line is the current trajectory of new reported cases per day in Ontario if the current level of non-pharmaceutical interventions are maintained. The pink line shows the possible trajectory under vaccine roll-out of equation (8) and non-pharmaceutical interventions and maintained without relaxation.

#### 3.2.2 Relaxation

For the sake of comparison, we present a scenario whereby NPIs relaxation starts on December 8, 2020 without any vaccination plan. This scenario is presented in Figure 5 as a worst case scenario stemming from the model. As expected, without any mitigation strategies we would observe a greater number of infected cases, compared to the case where a 75% vaccination rate is assumed with NPIs remaining in place at the current levels of acceptance and adherence.

**Figure 5:**
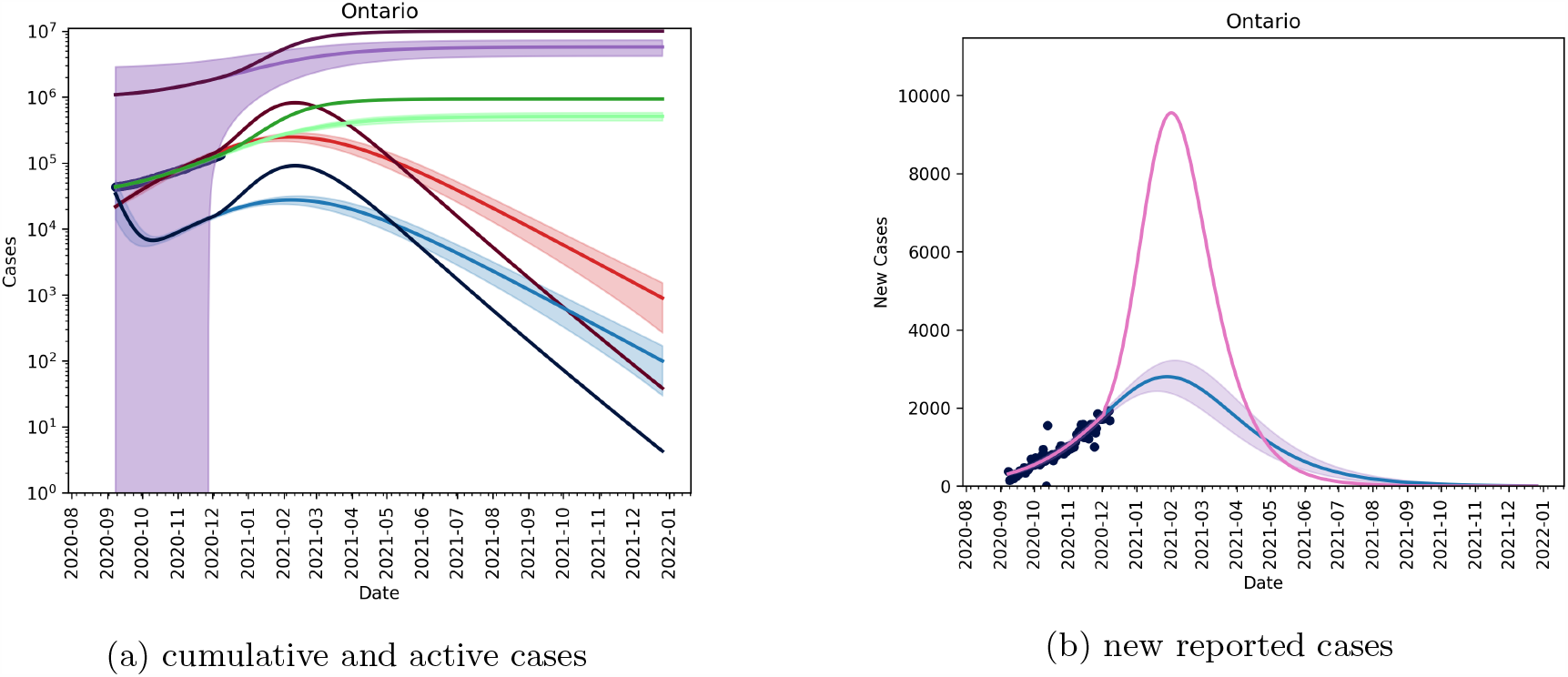
Removing non-pharmaceutical interventions on December 8, 2020 with no re-implementation and without vaccination. Again, blue line is the current trajectory, shaded region is the 95% confidence area and the pink line is the modified scenario.

The figure shows that by removing NPIs without any vaccination, we will see an approximately 80% attack rate in Ontario. This is in line with estimates provided by other models []. cumulative and active cases (b) new reported cases

#### 3.2.3 Assuming a 75% vaccination rate in Ontario and NPIs relaxed quickly

Figure 6 shows the effects of relaxing NPIs too early. For this scenario we set *T* = 14.5 and *θ* = 10. We see that all active cases are still resolved by September, 2021 (Figure 6a) and we see no new cases by July 2021 (Figure 6b), but the in the interim the caseload in the province increases.

**Figure 6:**
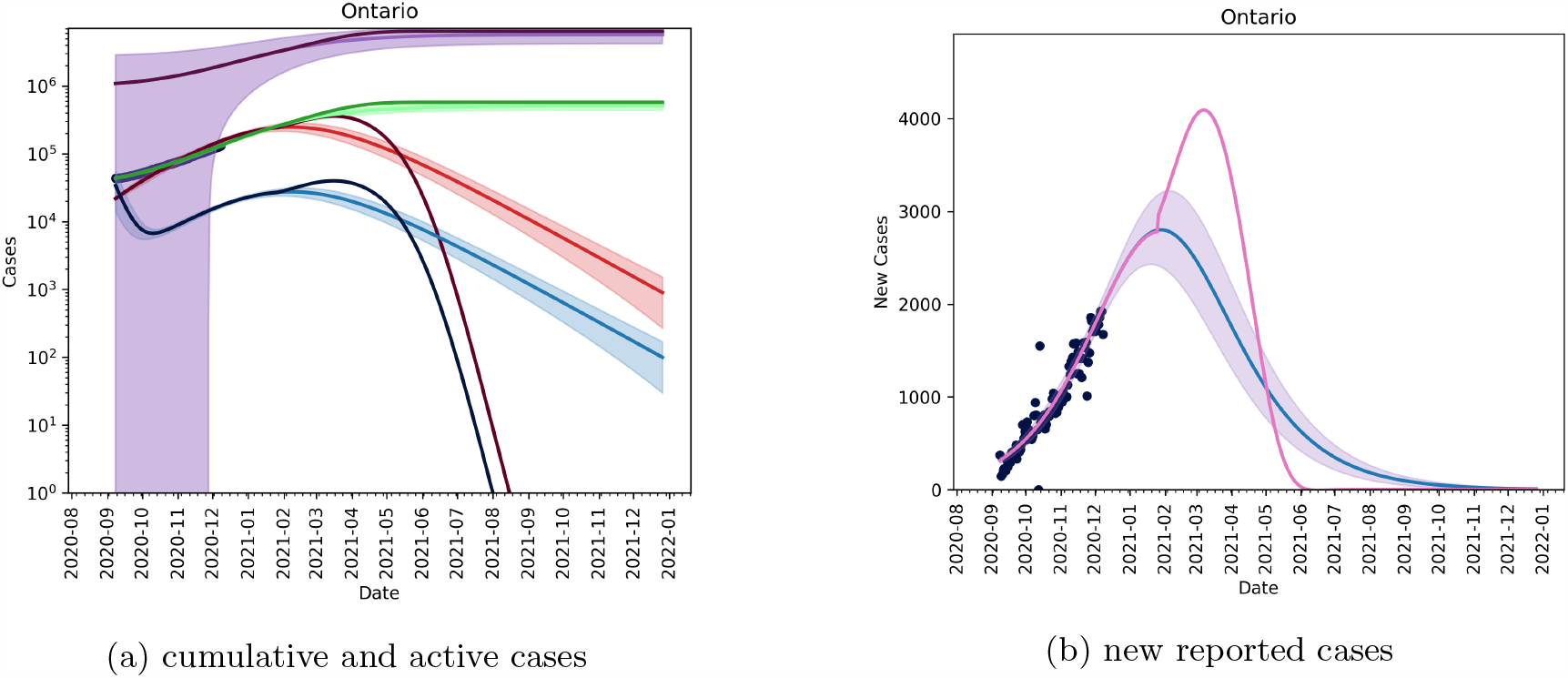
Removing non-pharmaceutical interventions on January 31, 2021 creates a massive increase in cases, but vaccine roll-out has a large effect on mitigating cases.

#### 3.2.4 Assuming a 75% vaccination rate in Ontario current NPIs relaxed from March 31, 2021

Figure 7 shows that phasing in relaxation starting on March 31, 2021 allows the province of Ontario to maintain control of vaccination roll-out and reopening. In this case, we vary *θ* and set *T* = 20.4. We seen in Figures 7a and 7d that if we phase-in reopening slowly then we can maintain control of the outbreak and allow for the realization of vaccination to be seen. Figures 7b, 7e, 7f and 7c show that increasing the speed of reopening can be detrimental to the control of the outbreak and can outpace the vaccine roll-out.

**Figure 7:**
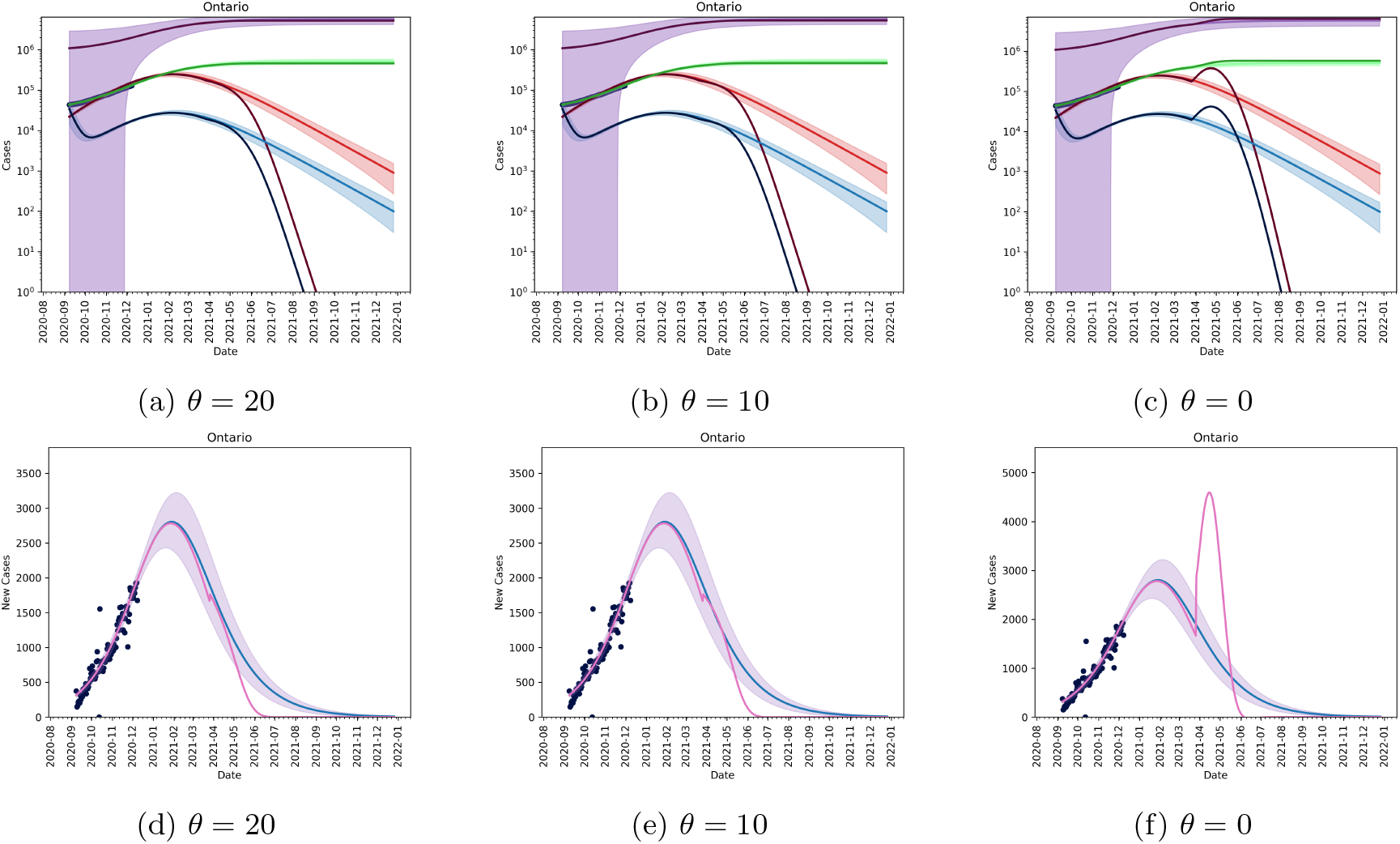
Removing non-pharmaceutical interventions on March 31, 2021 can be controlled.

#### 3.2.5 Assuming a 75% vaccination rate in Ontario current NPIs completely removed on May 01, 2021

Figure 8 shows the effects of removing all restrictions, immediately (*θ* = 0) on May 1, 2021. We see that by May 2021, we are able to reopen without risk of significant increase of new cases. In Figure 8a we see that the cumulative caseload does not change significantly, but Figure 8b shows a non-negligible increase in new cases as reopening occurs.

**Figure 8:**
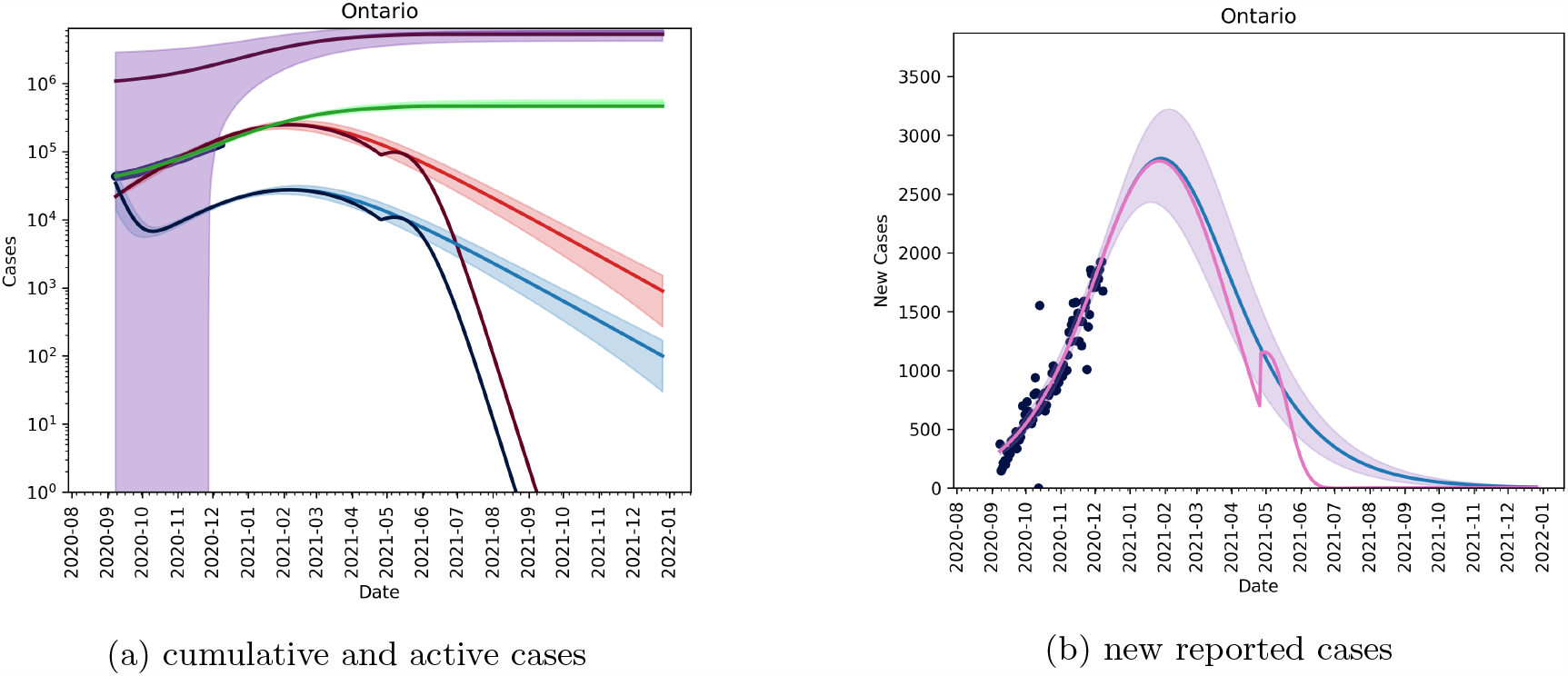
Removing non-pharmaceutical interventions on May 1, 2021; we need to be fairly low on the outbreak curve to remove restrictions without phasing.

### 3.3 A linear vaccination strategy

We can also look at a more hypothetical, but equally valid vaccination strategy employed by the equation

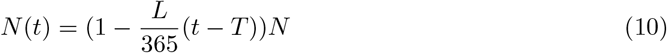

which suggests a constant vaccination rate across the population. Figure 9a we see that active cases with such a strategy are not resolved until December 2021, and in Figure 9b we see that we continue to observe new cases until August 2021.

**Figure 9:**
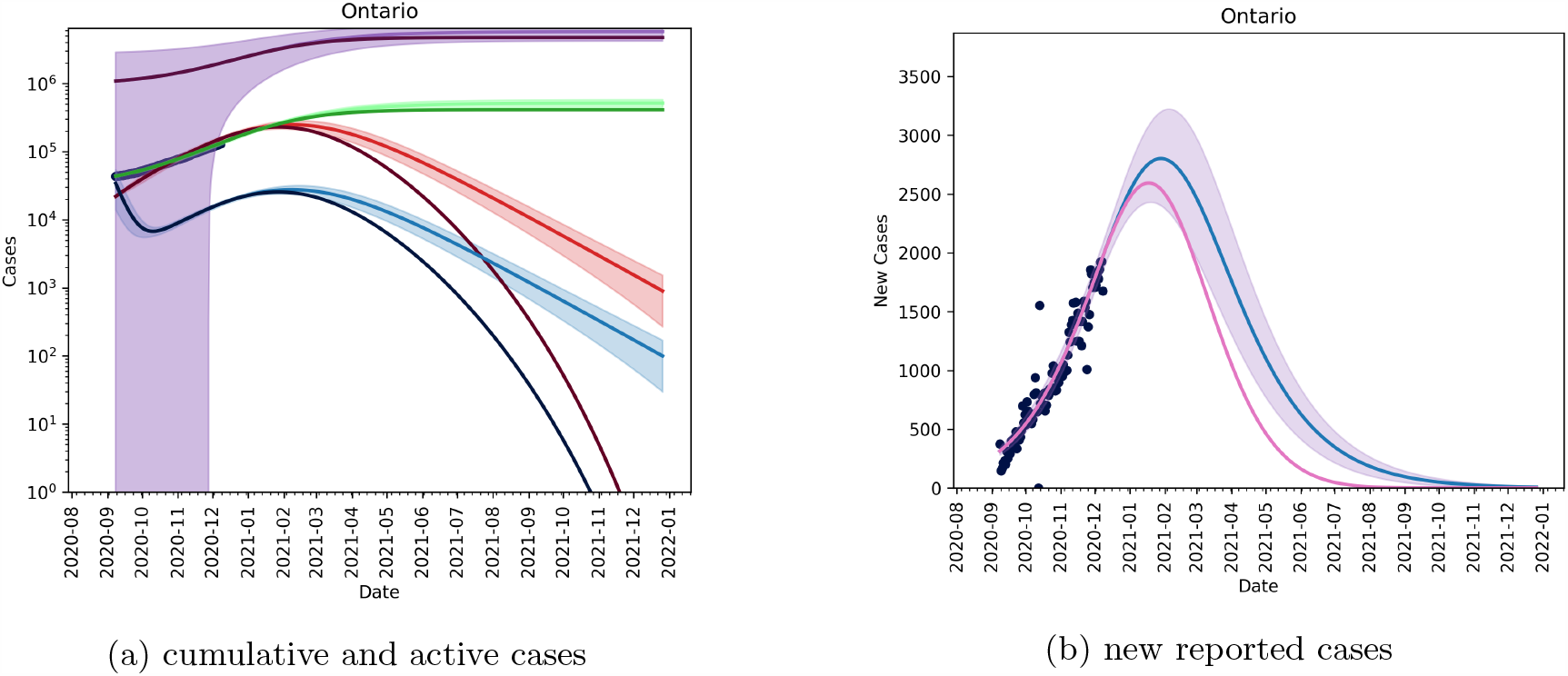
A constant vaccination rate will allow for a mitigating outbreak peak at the expense of a longer outbreak tail.

### 3.4 Date the effective reproduction number will be less than 1 for different vaccination % and NPIs relaxation measures

The effective reproduction number is given by

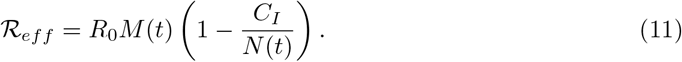

It shows the average number of cases one active case will create.

Figure 10 shows the effects of the different relaxation/vaccination strategies on the effective reproduction number. The different coloured lines represent the different scenarios described above. The dashed lines show when we have an *R*_*eff*_ < 1 for each strategy. The figure suggests that reopening in May 2021 presents the least risk for a new outbreak to occur. By reopening earlier, we risk *R*_*eff*_ increasing above 1 for a longer period of time, creating increased risk for large outbreaks.

**Figure 10:**
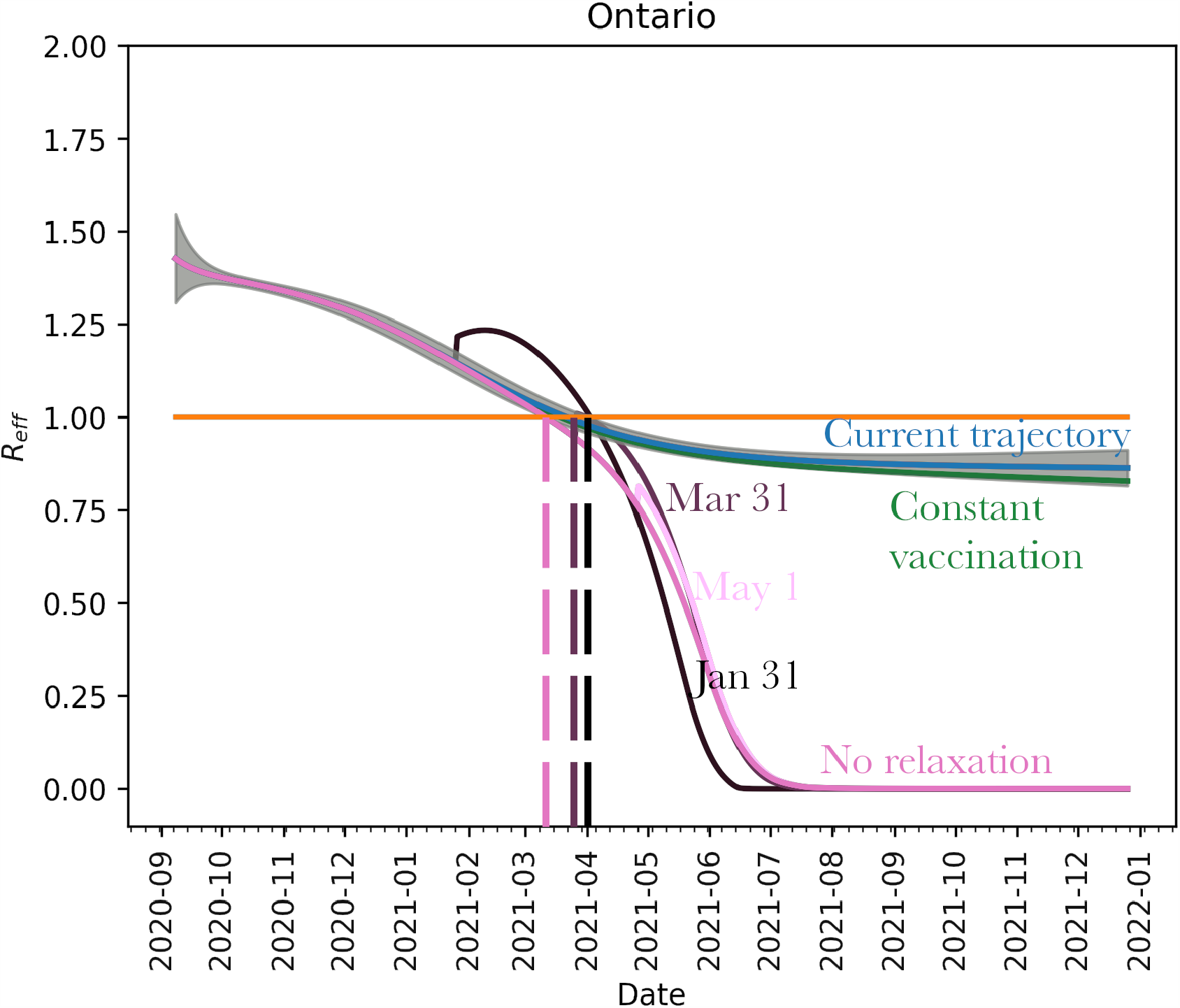
A reduction of ℛ_*eff*_ with target dates for different scenarios described above. We see that opening too early will cause ℛ_*eff*_ to increase above 1 which will increase the risk of localized outbreaks. If relaxation is delayed until May 2021 and phased-in, reopening can occur with very little risk.

## 4 Interpretation

We explored the effects of an integrated/combined package of both pharmaceutical and non-pharmaceutical public health control measures. We demonstrated that these two interventions need to be fully harmonized in order to be able to properly control the spread of the outbreak. In the literature, there are some models modelling the impact of COVID-19 vaccination in terms of the epidemiological trend of the pandemic. For example, Libotte and coauthors [10] designed a mathematical compartmental SIR model and explored two major scenarios, according to the optimal control theory, and informing the model by real-data from China. The first strategy assessed consisted in minimizing the number of infected individuals, whereas in the second scenario also the minimization of vaccine dose was taken into account.

Jentsch and colleagues [11] devised a coupled social-epidemiological model, simulating schools and unessential workplaces closures, utilizing an evolutionary game theory-based approach and exploiting mobility data. Some vaccination strategies were age-based, whereas others targeted social contacts, or particularly frail and vulnerable groups. Immunization could curb deaths by 22 to 77%. Vaccine availability impacted the proportion of preventable deaths: in case of vaccines already available since January 2021, targeting seniors first would be the most optimal strategy, whereas, in case of availability from July 2021 on, targeting social contacts first would result into a better control of the outbreak.

Similar results were obtained by Grundel and coworkers [12], who combined an optimization-based control approach with an age-dependent epidemiological model. To be able to relax physical/social distancing measures, a vaccination strategy targeting high-risk subjects was more effective in the short-term, whereas an approach based on the reduction in social contact rates was the optimal choice in the long-term. In terms of vaccine doses and effectiveness, authors found that greater amounts of a less effective vaccine resulted in more relaxed NPIs with respect to less amounts of a more effective vaccine.

We are able to leverage the current outbreak trajectory through data-fitting to refine scenario-based approaches that compete reopening with vaccination. Current vaccination strategies employed throughout the country are focused on high-risk rather than high-transmission populations. As reopening is, generally, not age-dependent (except school re-openings), and the focus of vaccination is on reducing risk, not necessarily transmission, the questions addressed herein would not benefit from an age-structured model. The goal is to give potential dates to consider reopening with lowest risk, and timelines of potential herd immunity (in most cases, our results estimate that we will reach minimal cases per day by the end of second quarter 2021). While an age-structured model would be beneficial for developing policies in greater detail, the age-structured case data is sparse at best. In [13], age-related data is given at a weekly resolution and is updated infrequently. While the model and methods introduced in [6] and used here are able to fit age-structured data (even those sparsely provided) it is difficult to extend scenarios from such data at said resolution.

Moreover, our study focused at the province level (Ontario), as a case study. At the global level, few predictive models exist. For instance, by incorporating expert opinions and mathematical models, McDonnell et al.[14] have estimated that due to the time of manufacture and distribution of the vaccine, sufficient doses globally are likely not to be available before September 2023. Whilst our model equips local decision- and policy-makers with projections about the future epidemiological trend of the COVID-19 outbreak. It would be extremely useful to provide public health agencies across the world with a tool that is flexible enough to explore the effect of different mitigation strategies.

## Conclusion

Re-opening and vaccine deployment is a careful balancing act. Using our mathematical model, we extensively explore the effect of adopting various vaccination and relaxation strategies on the COVID-19 epidemiological long-term projections. Our findings are able to provide public health bodies with important insights on the effect of adopting various mitigation strategies, thereby guiding them in the decision-making process.

## Data Availability

Data is publicly available.

https://www.canada.ca/en/public-health/services/diseases/2019-novel-coronavirus-infection.html#a1

